# Dynamic versus Continuous Interventions: Optimizing Lockdown Policies for COVID-19

**DOI:** 10.1101/2021.03.10.21253324

**Authors:** Kaan Akinci, Javier Fdez, Elena Peña-Tapia, Olaf Witkowski

## Abstract

In the context of the ongoing COVID-19 pandemic, while millions of people await the administration of a vaccine, social distancing remains the leading approach towards the effect commonly known as “flattening the curve” of infections. Over the last year, governmental administrations throughout the globe have implemented various lockdown policies in hopes of slowing down the transmission of the disease. However, the current lack of consensus on when and how these policies should be implemented reflects the need for further studies regarding these questions. In this paper, we tackle the issue of lockdown policy management, in particular in terms of lockdown placement (how often, when, and how long these periods should be), in order to minimize the peak of infections in a specific population. We introduce a novel combination of classic mathematical disease modelling using the equation-based SEIR model, and Evolutionary Strategies (ES) for optimizing the peak of infections. The method is evaluated using data collected in different countries, and a particular focus is placed on the study of the effect of specific model parameters on lockdown optimization, such as the transmission rate (*β*), of which 4 alternative modelling functions have been proposed and analyzed. Our results indicate that this transmission rate parameter significantly influences the resulting optimal strategies. In particular, the presence of a gradual decay of the rate of transmission during lockdown leads to longer, more sparsely placed confinement periods while an abrupt, instantaneous drop in the amount of contacts per person favors shorter but more frequent lockdowns. Although these results are limited by the scope of action provided by the simplicity of the SEIR model, they suggest that the influence of the evolution of the rate of transmission along the disease should be assessed in further studies with alternative optimization strategies (agent-based) and models (SEIRSHUD).

## Introduction

The SARS-Cov2 virus, first reported in China at the end of 2019, has taken the world by storm: triggering a global healthcare crisis, curtailing international mobility, and severely impacting financial markets and the global economy. As of March 1st, 2021, the numbers of global infections and casualties stand at 114.7 million and 2.6 million, respectively, and have kept increasing on a daily basis since then ^1^.

After an unprecedented global effort for fast vaccine development, we have started to see a light at the end of the tunnel. However, until vaccines have been safely administered to a large enough fraction of the world’s population, the main method for preventing the spread of contagion consists of reducing person-to-person contact, a strategy known as non-pharmaceutical intervention (NPI) (Eubank et al., 2020).

Since 2020, governments have implemented their own NPI policies, basing their decisions on a limited understanding of the disease and its contagious properties. Most leaders have opted for the most politically palatable trade-off between reducing the spread of infection and facing economic hardship. Some policies seem to have been more successful than others, but the complexity of the issue, and differences in social, economic, cultural and even immunological factors surrounding each individual country complicating the formation of a general conclusion. In other words, there is still a lack of consensus on when and how these policies should be implemented, reflecting the need for further studies regarding these fundamental questions. This lack of consensus motivates the work behind this paper.

Recent literature related to curbing the spread of COVID-19 covers two main types of interventions. On the one hand are *continuous lockdowns*, where the population is confined for several weeks, such as the ones enforced in China or Europe at the beginning of the pandemic. On the other hand are the so-called *dynamic lockdowns*, which are alternated intervals of strict and relaxed confinement periods. The effects of these interventions are often measured against each other or compared with softer mitigation measures.

One example is a study by Chowdhury et al. (2020), where the authors compared simulation scenarios with (1) no interventions, (2) consecutive cycles of mitigation measures followed by a relaxation period, and (3) consecutive cycles of suppression measures followed by a relaxation period. The analyses were based on a standard susceptible-exposed-infected-recovered (SEIR) compartmental model (Blackwood and Childs, 2018), and the resulting optimal strategy was the implementation of dynamic cycles of 50-day mitigation periods followed by 30-day relaxation intervals. Similarly, M. Kennedy et al. (2020) compare constant, intermittent, and “stepping-down” social distancing strategies using a susceptible-unsusceptible-exposed-infected-hospitalized-critical-dead-recovered (SUEIHCDR) model, which is an extension of the SEIR models using factors specific to COVID-19 ^2 3^. The authors concluded that a “stepping-down” (dynamic) policy was the best long-term social distancing strategy to minimize the peak number of active cases. In complementary work by Rawson et al. (2020) using an adaptation of the SEIR model, an optimizer was developed for determining the best strategy for lockdown release regulation to prevent the saturation of the health system. They concluded that the optimal solution would be a gradual approach: releasing half of the general population 2-4 weeks from the end of an initial infection peak, and waiting 3–4 months to allow for a second peak, before proceeding with the remainder of the population.

Overall, papers comparing different styles of interventions with SEIR-related models tend to point out dynamic strategies as the optimal to minimize the peak of active cases. However, these studies only compare pre-determined lockdown periods with specific fixed time frames. If we leave the task of lockdown placement and length assignment open for our algorithm to decide, would the results be consistent with previous findings?

The current go-to technique to study lockdown placement, used in the examples presented above, consists of modelling the infection curve using an epidemiological model and applying mathematical optimization methods to reduce the peak of the infection curve. The transmission dynamics of the virus follow a state-dependent structure, known as a Markov decision process, which can be exploited using reinforcement learning (RL) techniques. Indeed, several authors decided to try RL for lockdown policy optimization, since it can also schedule tasks in an adaptive manner (Glaubius et al., 2012). For example, Arango and Pelov (2020) used the Double Deep Q-Learning (Van Hasselt et al., 2016) algorithm to find optimum lockdowns policies for multiple lockdown lengths for the COVID-19 using an extended SEIR model. Similarly, Colas et al. (2020) built and demonstrated an environment for many Deep Learning algorithms that can optimize lockdowns for a variety of epidemiological models. Furthermore, Khadilkar et al. (2020) presented a quantitative way to compute lockdown decisions using RL for individual cities or regions that balances health and economic considerations. Lastly, a report by Miralles-Pechuán et al. (2020) proposed Deep Q-Learning and genetic algorithms to optimize the best sequences of actions governments can take to reduce the harmful effects of a pandemic and proved that their methodology is a valid tool to find actions that governments can take to reduce the adverse effects of a pandemic. Despite being a powerful tool when it comes to optimization of episodic optimization, RL presents its own challenges such as reward distribution (dense or sparse), number of episodes, and difficulty of parallel computing (Salimans et al., 2017). A competitive alternative to the use of reinforcement learning for lockdown policy optimization are evolutionary algorithms. They are easily parallelizable, and their loss functions are simple to model. Besides, due to their population-based structure, these algorithms yield sub-optimal solutions in addition to optimal ones, which is helpful for understanding the dynamics of the lockdowns and the solution space. In the current literature related to lockdown optimization, most researchers have implemented Genetic Algorithms (GA), developed in the 1960s (Whitley, 1994) and inspired by Charles Darwin’s theory of natural evolution. These algorithms are commonly applied in computer science for optimization problems and reflect that the fittest individuals of the population are often the ones selected for reproduction in order to produce offspring for the next generation.

Pinto Neto et al. (2021) used a multi-objective GA design optimization on a epidemiological compartmental model, named SUEIHCDR, to compare scenarios related to strategy type, the extent of social distancing, time window, and personal protection levels on the transmission dynamics of COVID-19 in São Paulo, Brazil. Their results indicated that the ability to reduce social distancing depends on a 5–10% increase in the current percentage of people strictly following protective guidelines, highlighting the importance of protective behavior in controlling the pandemic. Furthermore, Miralles-Pechuán et al. (2020) implemented both RL and GA to determine the sequences of actions (confinement, self-isolation, two-meter distance or not taking restrictions) on a SEIR model according to a reward system focused on meeting two objectives: firstly, getting few people infected so that hospitals are not overwhelmed, and secondly, avoiding taking drastic measures that could cause serious damage to the economy. Interestingly, their approach based on RL outperformed the one based on GA. Another work by Zhang G (2021) combined GA and a long short-term memory (LSTM) neural network to optimize the infection rates on a susceptible-infected-quarantined-recovered (SIQR) epidemic spreading model, achieving good predictive ability on infection and death cases. However, after implementing and assessing such strategies in our experiments, we observed that the solutions found by the GA had high divergence, specifically for the runs where the rate of transmission varies over time. This shows how GA falls into local optima when increasing the complexity of the model.

Similar to GA, Evolutionary Strategies (ES) is an algorithm that uses a population of individuals that are evaluated by a fitness function and perturbed by mutation. It also has fewer hyperparameters compared to GA and does not suffer from settings with sparse rewards. After testing this algorithm, we observed that ES produced better and more robust results compared to GA, so we decided to use it for our experiment. Besides, to the best of the authors’ knowledge, this is one of the first studies to assess this algorithm for COVID-19 lockdown optimization.

Interestingly, there are few instances in recent literature that have used mathematical optimization methods to compare different interventions. One of the most relevant studies that developed a model to evaluate the spread of COVID-19 in South Africa under different policy scenarios is the one by Olivier et al. (2020). The scenarios analyzed how to flat-ten the curve to a level that the healthcare system could cope with, how to balance lives and livelihoods, and what impact the compliance of the population to the lockdown measures had on the spread of COVID-19. An interesting takeaway from this work is the implementation of variations in the rate of transmission throughout time, based on its dependency on factors such as social distancing, restrictions on travel, or working activity. The authors proposed the following function to model such variation over time:

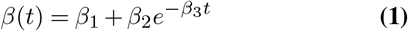

Motivated by the insights of this last work, we define the aim of this work as twofold. On one hand, we aim to expand the current insight on lockdown optimization strategies by experimenting with a combination of the SEIR equation-based model and a novel optimizer based on Evolutionary Strategies (ES). On the other hand, we aim to explore the influence of the transmission rate parameter (*β*) on the resulting strategy proposed by the optimizer. The motivation behind this second study lies in the difficulty of choosing the “right” constant transmission rate out of reported data present in datasets in work such as that by Max Roser and Hasell (2020). While most SEIR-based works choose a constant rate based on previous literature, figures 4 and 5 show evidence of a non-constant behavior. The introduction of an exponential decay-based (*β*) modelling by Olivier et al. (2020) inspired the 4 scenarios that will be proposed in a later section. While our findings focus on the particular case of the ES-based optimizer, the results are of relevance for all SEIR-based optimization methods.

**Figure 1.**
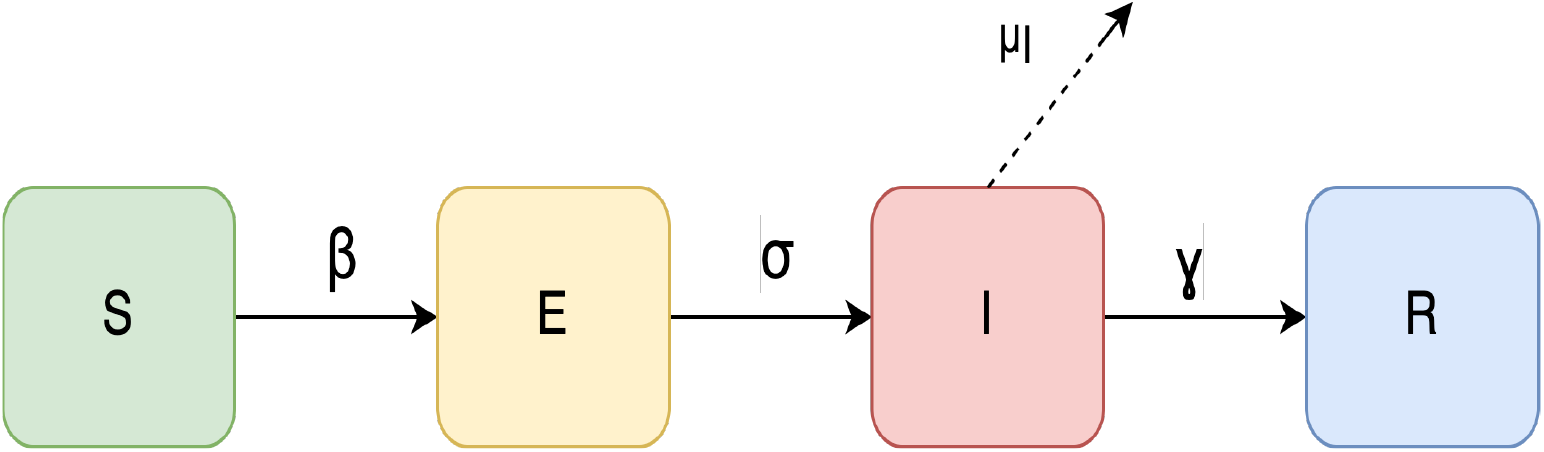
SEIR compartmental model.

**Figure 2.**
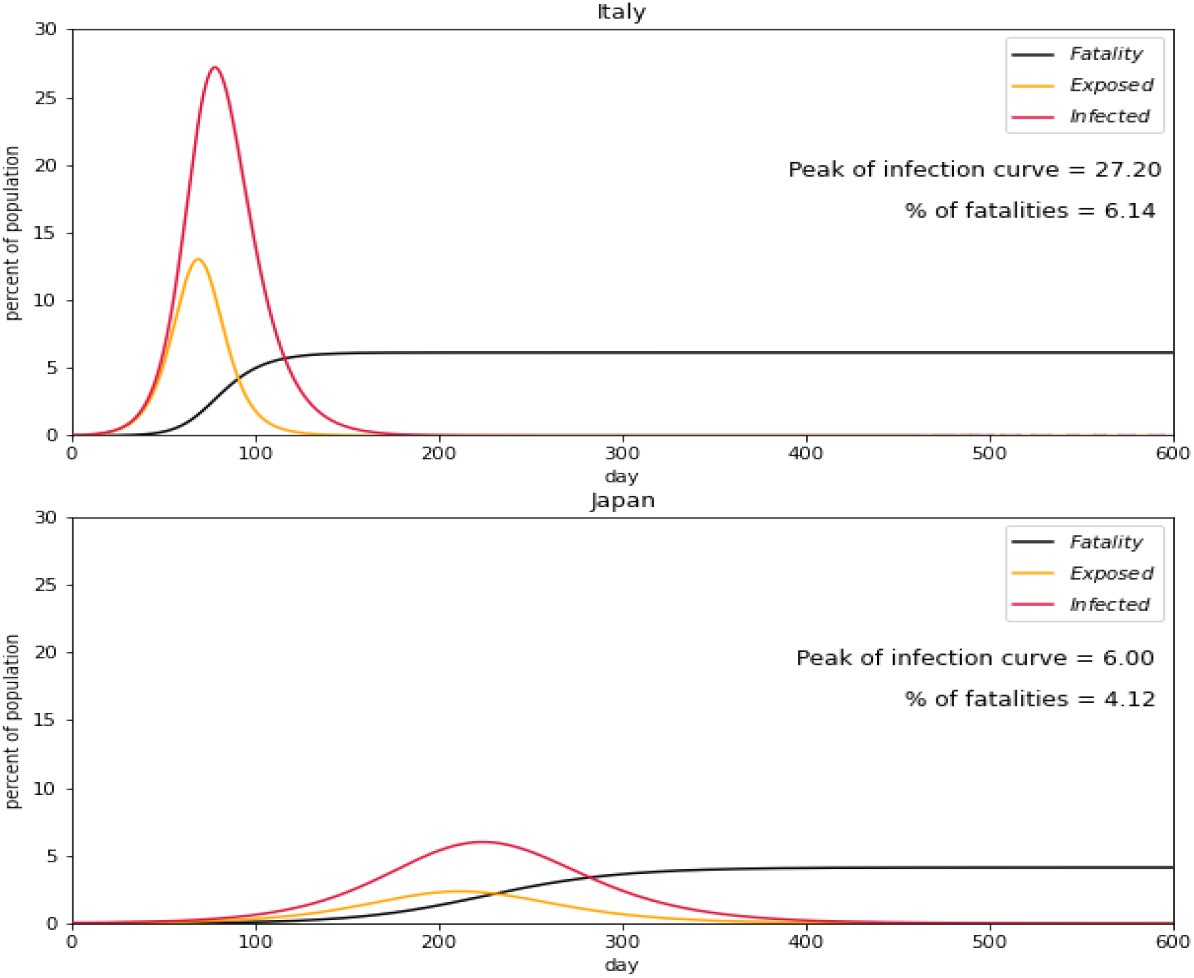
Simulation of the SEIR model for Italy (top) and Japan (bottom) with the input parameters of Table 1 and rate of transmission of 0.28572 for Italy and 0.12143 for Japan. The number of days can be seen on the x-axis, while the y-axis indicates the percentage of the affected population. The infected population is indicated by the red line, the exposed population is indicated by the yellow line, the population of those testing positive is indicated by the purple line, the black line indicates casualties.

**Figure 3.**
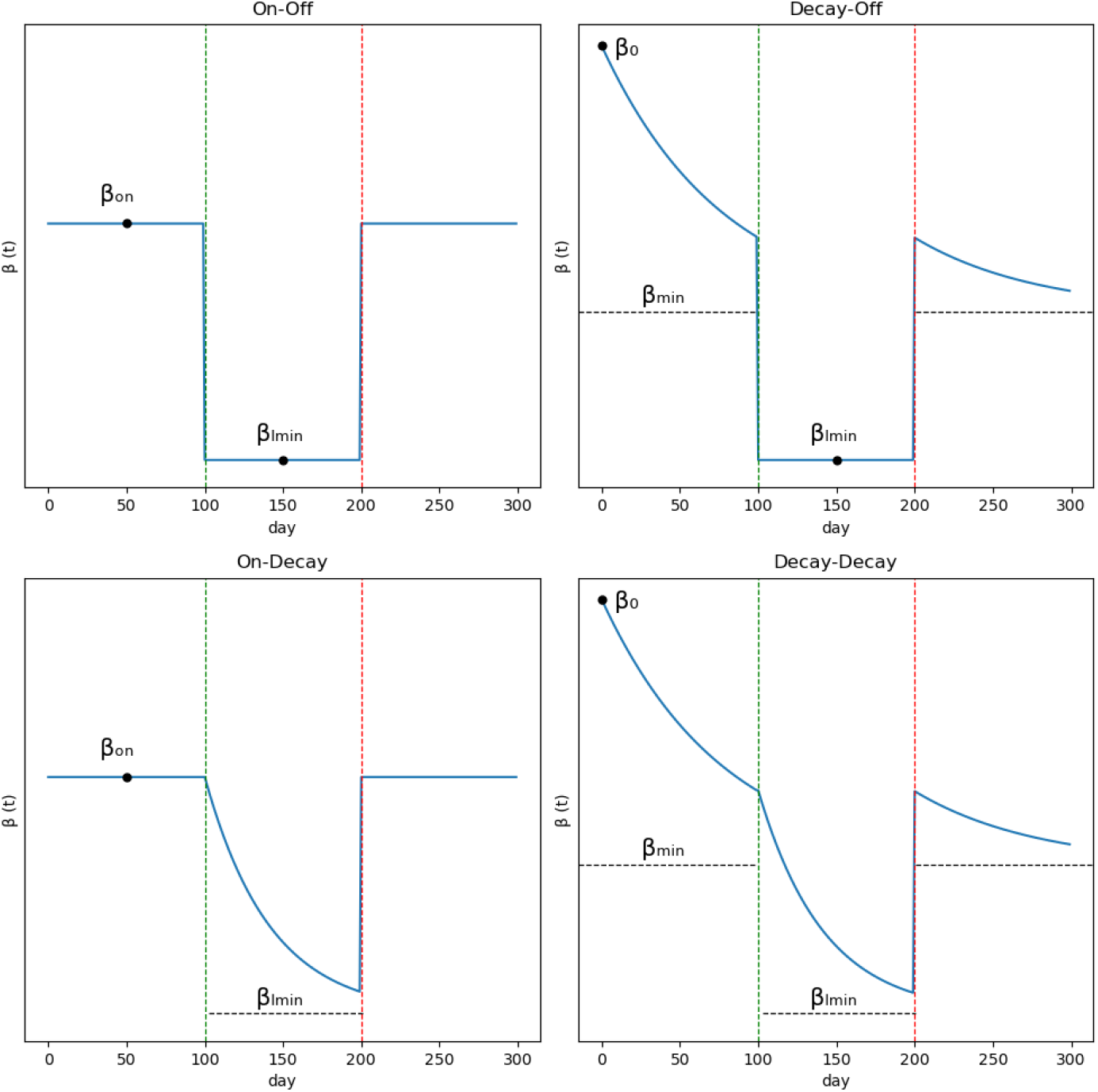
Scenarios for the different conditions of the beta decay.

**Figure 4.**
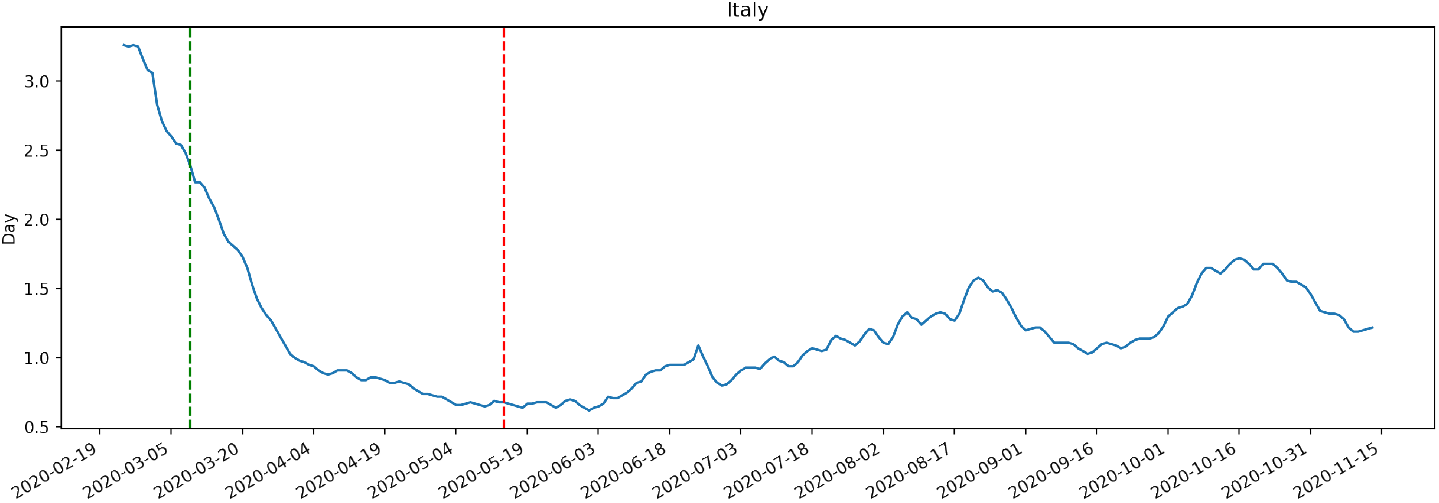
Rate of transmission in Italy. The green and red vertical lines indicate the beginning and end of the enforced quarantine.

**Figure 5.**
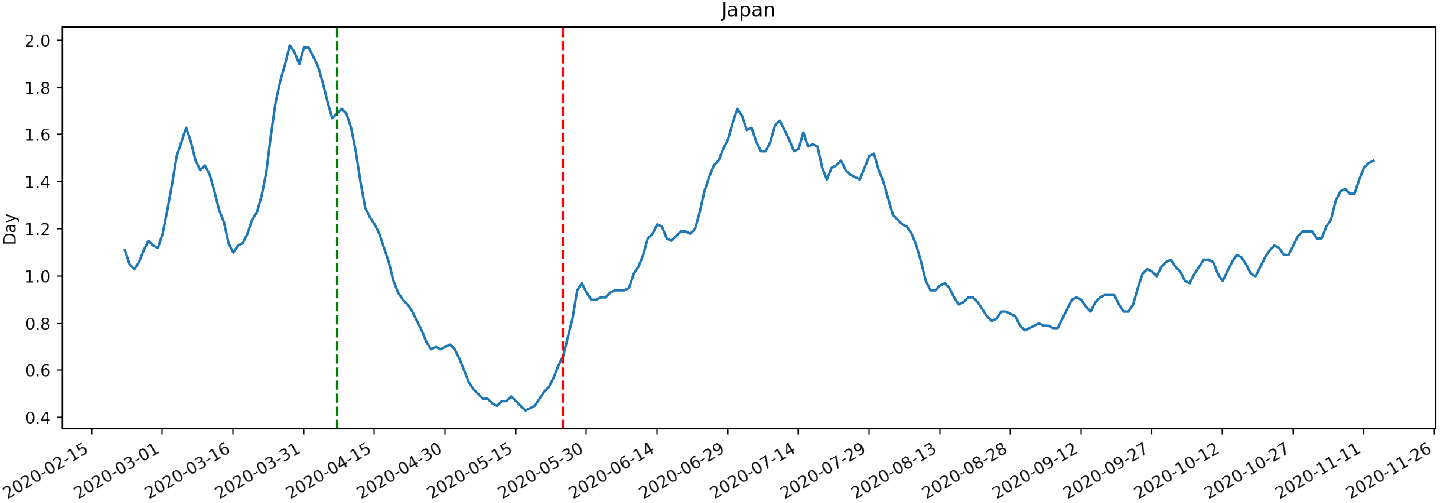
Rate of transmission in Japan over time. The green and red vertical lines indicate the beginning and end of the state of emergency.

The remainder of the paper is structured as follows: the Materials and Methods section provides a detailed description of our methodology and experimental setup, the results of which are discussed in the Results section, followed by our Conclusion and Future Work.

### A. Disclaimers

Before proceeding further, we wish to make explicit the limitations of this study. Firstly, the results presented were obtained through an SEIR model, an epidemiological model that characterizes a pandemic’s epidemic dynamics using ordinary differential equations. Secondly, our results are based on an estimated set of epidemiological parameters, which may not accurately fit the real data. Furthermore, the estimated values for the rate of transmission are based on the data reported by governments, which may be inaccurate due to the lack of tests at the beginning of the disease. Given these limitations, further research is required before applying these findings to real-world scenarios.

## Materials and methods

This section presents the methodology of our study for lockdown policy optimization, where we have tried to minimize the peak of infections using an ES optimizer for the SEIR model with 4 alternative lockdown periods summarized in Table 5, the fixed parameters mentioned in Table 1, and the four different modelling functions for the rate of transmision (*β*) from Table 2. With the aim of achieving greater generality in our conclusions, these simulations have been carried out with data from two very different countries (Italy and Japan), leading to a total of 4×4×2= 32 scenarios.

**Table 1.** SEIR parameters.

| Parameter | Description | Value | Reference |
| --- | --- | --- | --- |
| $\sigma$ | Rate of progression (inverse of the incubation period) | 0.2 | Linton et al. (2020), Rawson et al. (2020) |
| $\gamma$ | Rate of recovery (inverse of the infectious period) | 0.07142 | Jung et al. (2020) |
| $\mu_i$ | Rate of mortality from the disease (deaths per infectious individual per time) | 0.004785 | Rawson et al. (2020) |
| $init_N$ | Total number of individuals | 1000000 | |
| $init_E$ | Initial number of exposed individuals | 0 | |
| $init_I$ | Initial number of infectious individuals | 500 | |

**Table 2.** Type of decay for the rate of transmission

| Named | Type of decay |
| --- | --- |
| On-Off | No decay neither within or outside the lockdown |
| Decay-Off | Decay outside the lockdown |
| On-Decay | Decay within the lockdown |
| Decay-Decay | Decay within and outside the lockdown |

**Table 5.**
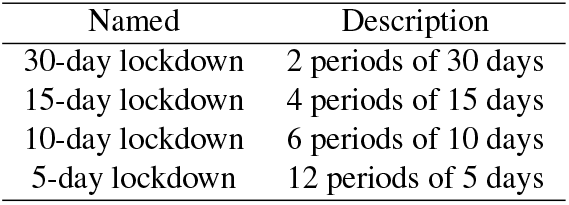
Type of lockdown policies.

The structure of this section is as follows: (A) It begins with an explanation of the SEIR equation-based model, followed by (B) the choice of model parameters, (C) different lockdown scenarios subject to study, and finally (D) an explanation of the optimization algorithm based on evolutionary strategies.

### A. Equation-based model

There are two main modelling alternatives for the simulation of viral diseases at a pandemic level: equation-based models (EBM) and agent-based models (ABM) (Sukumar and Nutaro, 2012). The former (EBM) defines a set of differential equations and focuses on efficiently capturing global interactions, ignoring local phenomena such as demographic characteristics, movement patterns, or differences in exposure levels for different subjects. The latter (ABM), models the disease as a collection of autonomous decision-making entities called agents. While agent-based models can offer interesting insights about the effects of specific agent interactions, when studying large scale behaviours they can suffer from a rapid increase in complexity. Any addition of or change in a small parameter may drastically change the overall behaviour of the simulation due to the inherent stochasticity of the system, increasing the computational cost and complicating the task of the optimizer. In our study, we have decided to use an EBM model because (1) it is deterministic, leading onto more stable solutions which simplify the posterior analysis, and (2) less computationally expensive than ABM models.

#### A.1. SEIR model

Specifically, we have developed a Susceptible-Exposed-Infected-Recovered (SEIR) compartmental model (see Figure 1), where *S* defines the fraction of susceptible individuals in a population (those able to contract the disease), *E* indicates the fraction of exposed individuals (those who have been infected but are not yet infectious), *I* specifies the fraction of infected individuals (those infected and capable of transmitting the disease) and *R* is the fraction of recovered individuals (those who have become immune).

This model can be augmented with interesting variables such as rate of re-susceptibility (inverse of temporary immunity period) (Bjørnstad et al., 2020) or hospitalization capacity (Veloz et al., 2020), present in related papers. However, given the difficulty of accessing to reliable data to provide a rigorous analysis of these variables, we have decided to simplify our scope to the basic SEIR model and leave the introduction of further variables for future works.

The equations of the SEIR model are:

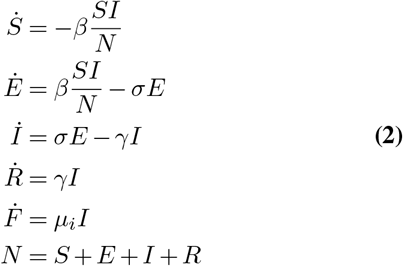

The transmission of the disease takes place when a susceptible individual (S) comes into contact with an infected individual (I), and is represented in the first equation of the system. This interaction is parametrized by the rate of transmission, *β*, which represents the potential number of transmissions per susceptible-infectious individual. The progression of the disease represents the change of category from exposed (E) to infected (I), and is parametrized by the rate of progression, *σ*, which is conversely the inverse of the incubation period of the disease (time passed from exposure to infection). The two remaining parameters are the rate of recovery, *γ*, which is the inverse of the infectious period, and the rate of mortality, *µ*_*i*_, which represents the deaths per infectious individual per time unit.

In order to perform simulations with this model, an additional set of parameters is required to set the initial conditions of the model variables. The parameter *init*_*N*_ indicates the total number of individuals, the parameter *init*_*E*_ the initial number of exposed individuals, and *init*_*I*_ the initial number of infectious individuals.

#### A.2. SEIR parameter selection

An adequate parameter selection {*β, σ, γ, µ*_*i*_} is key for the success of the SEIR predictions, and also one of the most challenging aspects of epidemiological modelling. For the set {*σ, γ, µ*_*i*_}, we have opted to adopt representative constant values agreed upon in a series of studies available in recent literature, gathered in Table

1. This table also includes the initial simulation values for the variables S, E and I.

However, this general consensus does not seem to apply to the rate of transmission {*β*}, where current available data reveals different values for different countries in different points in time. Indeed, some authors (Olivier and Craig, 2020) have proposed that the value of the rate of transmission reaches a peak at the beginning of the pandemic wave, and gradually decreases following an exponential decay function, unless interrupted by lockdown periods, which force *β* = 0. We have decided to provide a detailed study for the interaction of this parameter and its variation over time with the lockdown optimization task. For this purpose, we have gathered data from two countries with very different COVID transmission patterns: Italy and Japan. Section A.3 dives deeper into the modelling of rate of transmission for this work.

In addition to the parameters summarized in Table 1, running a simulation requires the definition of the total length of the action period. Following current available data on the evolution of the COVID pandemic, and under the assumption that vaccination efforts are being carried out in parallel to lockdown policies, a reasonable simulation time ranges from the beginning of the disease, at the end of 2019 (Zhou et al. (2020) reported the 19th of December, 2019), to mid 2021, with lockdown policy implementations starting around March 2020. This leads to a total simulation period of 600 days, with an action window of 480 days.

Figure 2 shows the results of a baseline simulation, which corresponds to the spread of the disease in Italy and Japan during the period of study without any intervention. The only difference between both models in the value of the transmission rate {*β*}, set to 0.171 for Italy and 0.121 for Japan. In the absence of lockdowns, the peak of the infection curve reaches approximately 27.20% and 6.00%, and fatality rate at the end of the disease is of 6.14% and 4.12% of the total population for Italy and Japan, respectively.

#### A.3. Rate of transmission

As introduced in the previous section, the rate of transmission, *β*, defined as the potential number of transmissions per susceptible-infectious individual, is difficult to characterize. However, it is most likely highly influential in the election of an optimal lockdown policy, as lockdown periods using the SEIR model are represented by reducing the rate of transmission to a value close to 0.

One of the novel contributions introduced by this paper is the study of different transmission rate characterizations in and out of lockdown, and their effects on a lockdown policy optimizer. Most works use a constant transmission rate extracted from the average values over a certain period of time (Chowdhury et al., 2020; M. Kennedy et al., 2020). However, a closer look into the current available data before any lockdown policy implementation such as in Italy (Figure 4) reveals a gradual decrease of the value of the rate of transmission from the moment the pandemic is declared until the first lockdown is implemented. Despite the quality of the data-taking process during these initial months adds an extra difficulty to the data analysis process as countries such as Japan, which did not resort to intensive testing, it is possible to observe a decay within the lockdown for both Italy and Japan (period between the green and red line for Figures 4 and 5). Olivier and Craig (2020) define this evolution of the transmission rate parameter in time with an exponential decay as following the next equation:

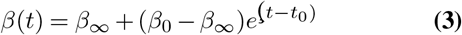

According to Olivier et al. (2020), this decay can be associated with factors such as the decrease on the number of susceptible individuals over time due to immunity, restrictions in travel, or an increase in social distancing. Instead, the existence of similar decay patterns during quarantine periods could be explained by an analogous reduction in susceptibility within the members of the same household.

As described in Table 2, we have considered the influence of the rate of transmission over time by setting four scenarios, where we study combinations of constant and variable functions in and out of lockdown. During the “decay” periods, the transmission rate *β*_*d*_(*t*) follows equation 3, while during the “no decay” periods, it remains at a constant value *β*_*nd*_. These combinations generate the four partial functions described in equations 4,5,6,7 and shown in Figure 3.

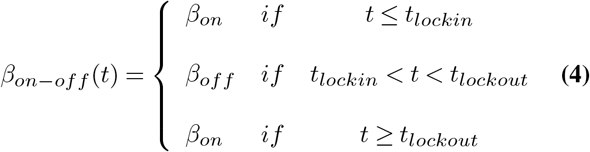

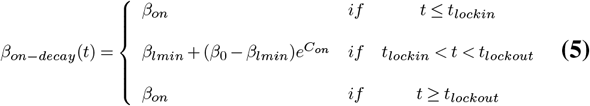

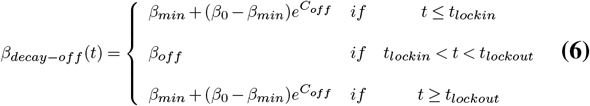

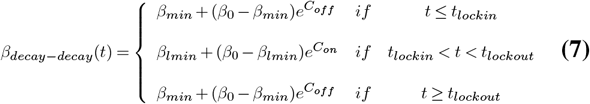

Where the meanings of each function parameters are:

1. *β*_0_: The rate of transmission at the beginning of the disease.
2. *β*_*on*_: The rate of transmission when there is no lockdown for the scenarios that there is no decay outside of the lockdown.
3. *β*^*lmin*^: The minimal value of the rate of transmission within the lockdown.
4. *β*_*min*_: The minimal value of the rate of transmission outside lockdown.
5. *C*_*off*_ : Exponential decay outside lockdown.
6. *C*_*on*_: Exponential decay within the lockdown.

The values of the initial and final transmission rates have been extracted from previous literature; these correspond to *β*_0_ (0.286 for Italy and 0.214 for Japan), and *β*_*min*_ (0.0179 for both countries). As for the remaining parameters, they result from fitting the 4 partial functions indicated in Table 4 to Italy and Japan’s transmission rate curves, extracted from the publicly available COVID-19 dataset maintained by Our World in Data, Max Roser and Hasell (2020) and gathered in Table 3.

**Table 3.**
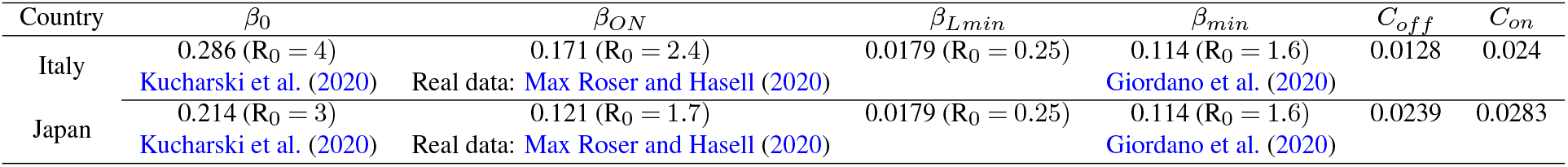
Japan and Italy’s parameters for the multiple scenarios.

| Country | $\beta_0$ | $\beta_{ON}$ | $\beta_{Lmin}$ | $\beta_{min}$ | $C_{off}$ | $C_{on}$ |
| --- | --- | --- | --- | --- | --- | --- |
| Italy | 0.286 ($R_0 = 4$) | 0.171 ($R_0 = 2.4$) | 0.0179 ($R_0 = 0.25$) | 0.114 ($R_0 = 1.6$) | 0.0128 | 0.024 |
|  | Kucharski et al. (2020) | Real data: Max Roser and Hasell (2020) |  | Giordano et al. (2020) |  |  |
| Japan | 0.214 ($R_0 = 3$) | 0.121 ($R_0 = 1.7$) | 0.0179 ($R_0 = 0.25$) | 0.114 ($R_0 = 1.6$) | 0.0239 | 0.0283 |
|  | Kucharski et al. (2020) | Real data: Max Roser and Hasell (2020) |  | Giordano et al. (2020) |  |  |

**Table 4.**
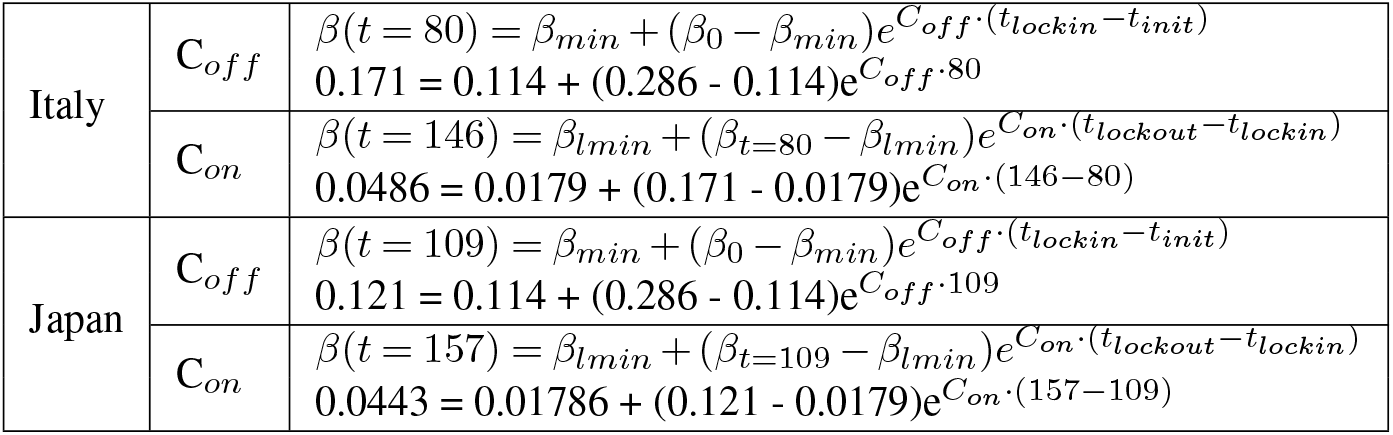
Formulas to obtain the decays of the rate of transmission within and outside the lockdown.

|  |  |  |
| --- | --- | --- |
| Italy | $C_{off}$ | $\beta(t=80) = \beta_{min} + (\beta_0 - \beta_{min})e^{C_{off} \cdot (t_{lockin} - t_{init})}$<br>$0.171 = 0.114 + (0.286 - 0.114)e^{C_{off} \cdot 80}$ |
| | $C_{on}$ | $\beta(t=146) = \beta_{lmin} + (\beta_{t=80} - \beta_{lmin})e^{C_{on} \cdot (t_{lockout} - t_{lockin})}$<br>$0.0486 = 0.0179 + (0.171 - 0.0179)e^{C_{on} \cdot (146-80)}$ |
| Japan | $C_{off}$ | $\beta(t=109) = \beta_{min} + (\beta_0 - \beta_{min})e^{C_{off} \cdot (t_{lockin} - t_{init})}$<br>$0.121 = 0.114 + (0.286 - 0.114)e^{C_{off} \cdot 109}$ |
| | $C_{on}$ | $\beta(t=157) = \beta_{lmin} + (\beta_{t=109} - \beta_{lmin})e^{C_{on} \cdot (t_{lockout} - t_{lockin})}$<br>$0.0443 = 0.01786 + (0.121 - 0.0179)e^{C_{on} \cdot (157-109)}$ |

Specifically, the parameters *C*_*off*_ and *C*_*on*_ were obtained using the starting time (green dashed line) and ending time (red dashed line) for Figures 4 and 5. With respect to Italy, the day when the lockdown measures were expanded to the entire country was March 9th, 2020. From that day, the government forbade any travel excluding the necessary for work and family emergencies. These measures lasted until May 14th, when stationery shops, bookshops and children clothing’s shops were allowed to open. In contrast, Japan did not impose a compulsory lockdown. Instead, it made a call for voluntary business closures which was widely respected. Nevertheless, Japan did impose severe border measures such as closing the international borders and not allowing foreign residents to reenter the country. Also, they enforced a mandatory quarantine after arrival and asked their citizens to wear a face mask in public. On April 7th, 2020, a one-month state of emergency was proclaimed for Tokyo and the prefectures of Kanagawa, Saitama, Chiba, Osaka, Hyogo, and Fukuoka. The lockdown was extended to the rest of the country on April 16th, 2020, for an indefinite period. The state of emergency was lifted for the whole country by May 25th.

### B. Types of lockdown policies

Once the model parameters have been established, in order to study the optimal lockdown policy it is necessary to set the principles of the lockdown strategy.

In literature, there are two main trends in the study of pandemic-related lockdown strategies. On one hand, there is the time-based strategy, where researchers set time-fixed consecutive cycles of lockdowns with a relaxation period between them, such as the studies by Chowdhury et al. (2020) and by M. Kennedy et al. (2020). On the other hand, there is the cost-based strategy, where a cost function is defined to model the trade-off between the economical costs of staying in lockdown and a peak in the number of infections. In these cases, the optimizer’s goal is to minimize this cost, and factors such as healthcare costs or impact of lockdowns on the country’s workforce are explicitly quantified. For example, Miralles-Pechuán et al. (2020) included health and economical costs, and Olivier et al. (2020) included a cumulative economic impact of lockdown in the loss function of its optimizer.

Because the quantification of the economical impact of lockdowns and health consequences derived from COVID-19 infections is a complex and nuanced matter, this work avoids the definition of an explicit cost function. Instead, we have opted for a mixed time-based strategy, where we set a limit on the total number of days that a country can remain in lockdown, and we divide it into several periods, while leaving the selection of when and how should this periods be assigned to minimize the peak of infection curve.

In particular, we observed that most of the countries implemented a lockdown at the beginning of the disease of approximately 60 days (2 months), so we decided to set as the total number of possible days for lockdown those 60 days, dividing them into four scenarios that differ in length and number, displayed in Table 5.

### C. Lockdown optimization algorithm

In order to find the optimal lockdowns time, we formulated the problem as a task scheduling problem. We decided to limit the length of the optimization period to 400 days and we distributed the lockdowns over those 400 days to minimize the peak of the infection curve. We also limited the number of lockdowns to observe the differences between long-term and short-term lockdowns, which is our resource in this problem’s context. Some of the main approaches found in the literature to solve such resource optimization problem in the context of COVID-19 are reinforcement learning (RL) and evolutionary algorithms. Because our experiment’s purpose is to find the optimal lockdown policy for the given situation rather than to find a generalised adaptive lockdown policy, we decided to avoid using reinforcement learning to avoid such issues. Compared to RL, evolutionary algorithms are easier to implement because the loss functions are easier to model and the algorithms are easy to train in parallel. Despite the fact that genetic algorithms (GA) are the most common evolutionary method (Pinto Neto et al., 2021; Miralles-Pechuán et al., 2020; Zhang G, 2021), we observed that they had high divergence caused by local optimums. Therefore, to obtain more reliable solutions, we decided to change our optimizer to Evolutionary Strategies (ES).

Similar to the GA, ES uses a population of individuals which are evaluated by a fitness function, and perturbed by mutation. Even though one of the most widely versions of ES is CMA-ES developed by Hansen and Ostermeier (2001), we decided to use a more nature-based implementation of ES called NES. This one was developed by Salimans et al. and is closely related to the work of Sehnke et al.. This model uses *θ* to set the distribution over the chromosomes, *P*_*ψ*_(*θ*) to represent our population and parameterized with *ψ*, and a fitness function calculated with *F* (*θ*). The goal is to find the optimal value for *ψ* that maximizes the average maximum value for 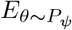. The algorithm uses gradient steps to update current population with the formula 8 (Salimans et al., 2017).

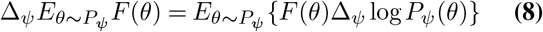

In this work we are searching for the optimal lockdown times, which are parameterized by *θ*. We initialize our population with Gaussian distribution with mean *ψ* and variance of *σ*^2^*I*. Because we defined our objective in terms of *θ*, we can use the formula 9 (Salimans et al., 2017).

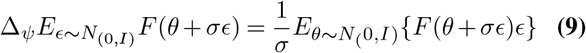

Then, we set the fitness function as:

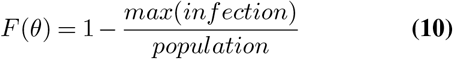

where *infection* is the infection curve in terms of number of people, and *population* is the number of people in the simulation. This corresponds to the percentage of *Susceptible* people at the highest point of the *infected curve*.

Because NES algorithm calculates the natural gradient in each step, it benefits from having a larger population size. However, computational cost also scales with the population size. In our work, we have experimented with different population sizes for the NES algorithm and found out that population size of 200 is sufficient for this optimization task. Once this population size was set, we experimented with different values for *σ* and *ψ*. In order to evaluate different solutions at the beginning and converge to the optimal one, we decided to change the *σ* and *ψ* parameters during the training and, as a strategy, we also decided to use simulated annealing (Rutenbar, 1989). Finally, we set *ψ* to be half of *σ* as in equation 11). In order to change the value for *σ* we used the formula 12.

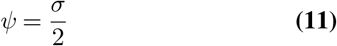

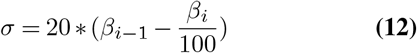

After experimenting with different values for *β*, we obtained the best results for *β*_0_ equal to 80.

## Results

The results will be analyzed from two different perspectives: on one side, we will try to answer to the question of lockdown placement, in particular, we will try to see if shorter and more frequent lockdowns (dynamic) are more efficient than longer lockdowns (continuous). In order to confidently answer this question, the results would need to be consistent for all proposed scenarios. The second perspective should help to shed light on this matter, as we will analyze the effects of the different transmission rate behaviors on the lockdown placement policies.

### A. Statistical analysis

This subsection provides an overview of the optimal solutions found by the proposed ES-based optimizer for placing a total of 60 days of lockdown in either 5-, 10-, 15- or 30-day long periods. The initial hypothesis, according to recent publications (Blackwood and Childs, 2018; M. Kennedy et al., 2020; Rawson et al., 2020), is that dynamic lockdowns are bound to perform better than longer, more sparse quarantine periods. As the optimizer is technically allowed to merge periods together, it might seem intuitive to only test the 5-day-long case. However, this configuration with redundant periods allows us to verify the consistency of the optimizer’s solution. For example, should 15-day lockdowns emerge as the leading solution, this would be shown both by a lower peak of infections of the 15-day case, and by a grouping of 5-day lockdowns in “packs” of 3. Figure 6 shows an example of one the 32 runs carried out during the experimental phase, in this particular case with data from Italy. Each quadrant corresponds to one of the scenarios presented in Table 2. At a first glance, we can confirm that all lockdown policies lead to a decrease in the peak of the infection curve, compared to the baseline simulation (Figure 2). Nevertheless, in practical cases it is important to remember that each day of lockdown has tremendous economical consequences to the region where it is implemented. Thereafter, we have assessed the results of the optimizer for each of the 32 proposed runs. Figure 7 reports the performance of the optimizer for each country, lockdown length, and rate of transmission modelling function. To evaluate the results, we have performed a three-way ANOVA applied to the peak of the infection curve, where the three factors were the (1) country, (2) type of lockdown, and (3) *β* function. The statistical results pointed out a significant interaction between the three factors (F(9, 128) = 22.0, p < 0.001), and the main effects of the country (F(1, 128) = 1143.7, p < 0.001), type of lockdown (F(1, 128) = 1537.2, p < 0.001), and type of decay (F(1, 128) = 48.3, p < 0.001). As a result, we carried out a two-way ANOVA test for both of the countries, Japan and Italy, separately.

**Figure 6.**
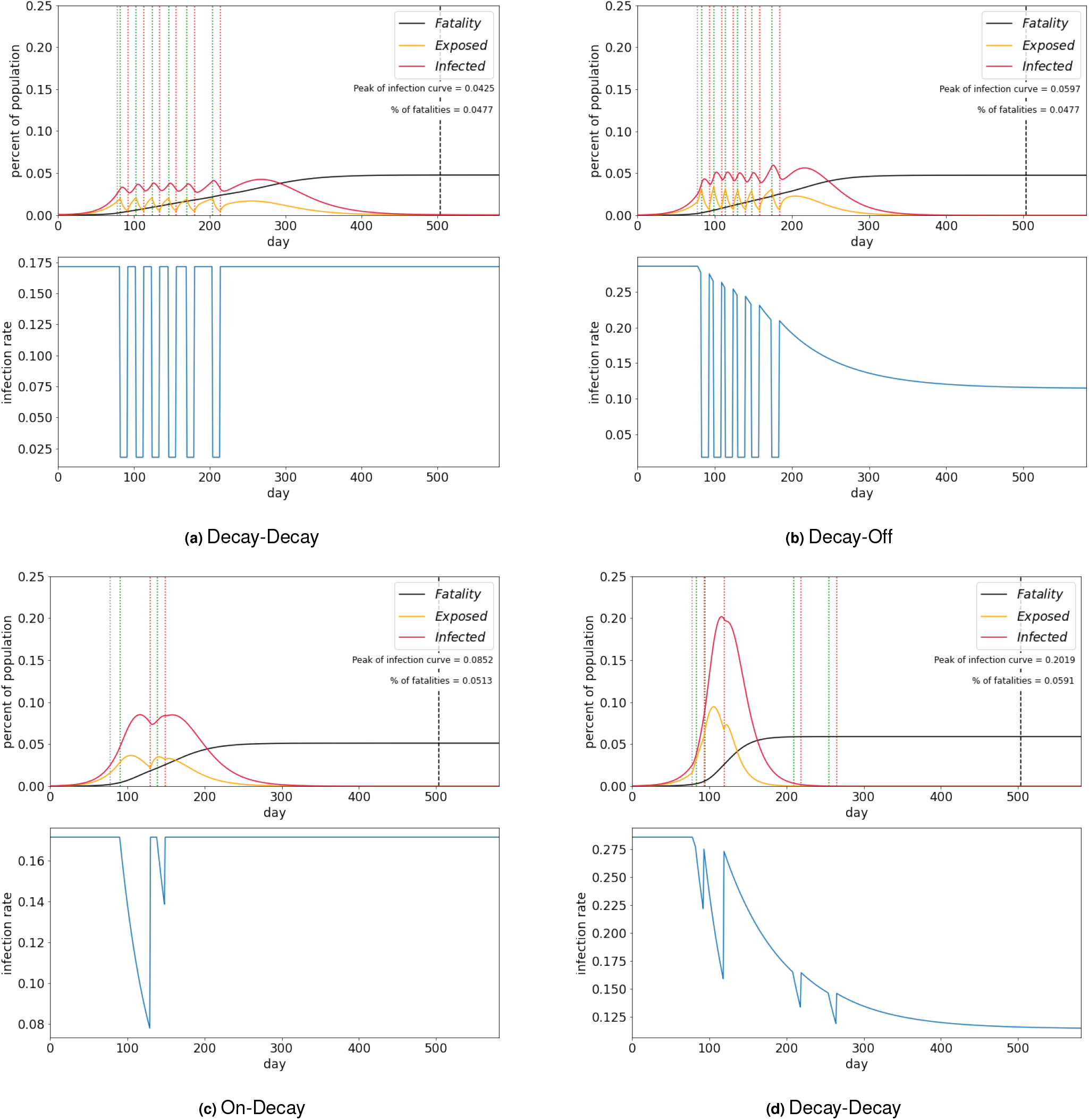
Italy 10-day’s run for the four rate of progression scenarios.

**Figure 7.**
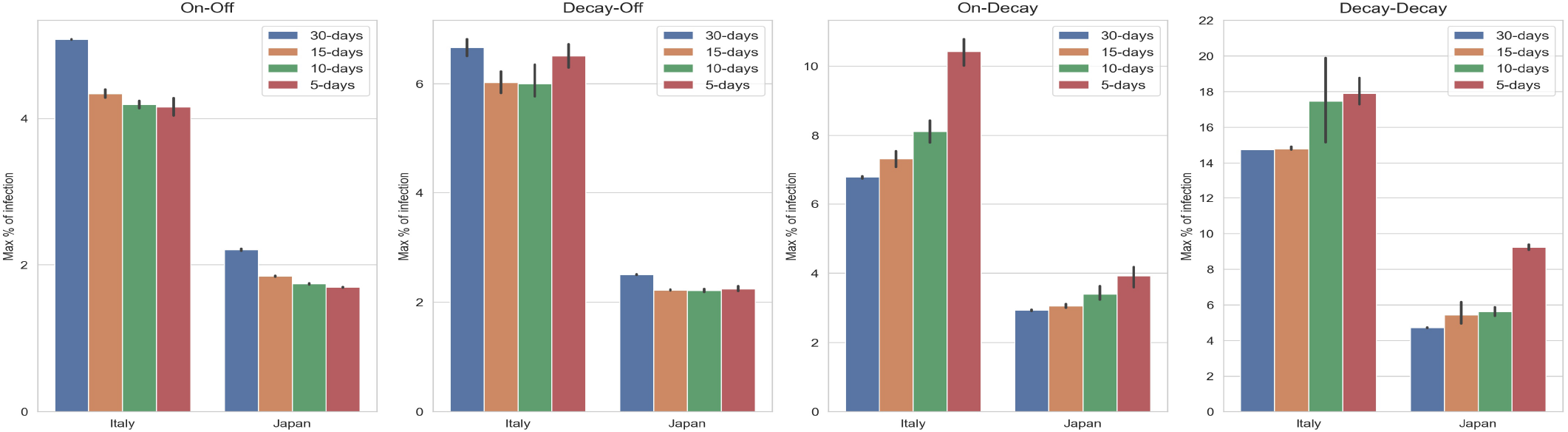
Overall performance of the runs for the SEIR simulation

For Italy, we found a significant interaction between the type of lockdown and *β* function (F(9, 64) = 53.8, p < 0.001). Hence, we implemented a one-way ANOVA for each of the *β* functions, showing a significant influence revealing effects as well between the type of lockdowns (On-Off: F(3, 16) = 1580.9, p < 0.001; On-Decay: F(3, 16) = 16.3, p < 0.001; Decay-Off: F(3, 16) = 80.0, p < 0.001; Decay-Decay: F(3, 16) = 92.4, p < 0.001). Therefore, we ran an independent t-tests between each condition of the type of lockdown and concluded that:

1. 5-day lockdown is statistically significantly better for On-Off
2. 30-day lockdown is statistically worse than the rest of the solutions for Decay-Off
3. 30-day lockdown is statistically better than the rest of the solutions for On-Decay
4. 15-day and 30-day are similar for Decay-Decay

The summary of these results is in Table 6.

**Table 6.** Optimal lockdown strategies found by the optimizer.

| Country | On-Off | Decay-Off | On-Decay | Decay-Decay |
| --- | --- | --- | --- | --- |
| Italy | 5-day | 5-day, 10-day, 15-day | 30-day | 15-day, 30-day |
| Japan | 5-day, 10-day | 10-day, 15-day | 30-day | 15-day, 30-day |

For Japan, the statistical analyses were very similar, showing as well an interaction between the type of lockdown and *β* function (F(9, 64) = 46.4, p < 0.001). Subsequently, we implemented a one-way ANOVA for each of the *β* functions, which revealed that there was an effect of the type of lockdowns (On-Off: F(3, 16) = 107.0, p < 0.001, On-Decay: F(3, 16) = 107.1, p < 0.001, Decay-Off: F(3, 16) = 6.83, p < 0.01, Decay-Decay: F(3, 16) = 6.06, p < 0.01). Hence, we ran independent t-tests between each of the conditions of the type of lockdown. Results indicated that:

1. 5-day and 10-day lockdowns are the optimal solutions for On-Off
2. 10-day and 15-day lockdowns are the optimal solutions for Decay-Off
3. 30-day lockdown is statistically better than the rest of the solutions for On-Decay
4. 15-day and 30-day are similar for Decay-Decay

The summary of these results is shown as well in Table 6.

### B. Discussion

In a nutshell, this initial evaluation lead to interesting findings: all factors of study have a significant impact on the peak of infections, but it is still possible to identify generalized trends throughout the results. Whenever an exponential decay function is applied to the rate of transmission inside the lockdown (cases on-decay and decay-decay), the optimizer favors longer lockdowns, contradicting our initial hypothesis. On the contrary, when this transmission rate inside lockdown remains constant, the optimizer behaves as expected and shows better performance with shorter lockdowns following a dynamic strategy. These trends are consistent across countries, despite the differences in the absolute value of the peak of infections.

Interestingly, in the cases where longer lockdowns are favored, the optimizer response is not consistent, as it does not succeed in grouping shorter periods to form longer lockdowns (for example, in the decay-decay graphs from Italy or Japan). The most likely explanation for this phenomenon is a fault of the optimizer when dealing with shorter lockdown periods, where the introduction of the internal exponential decay function hinders its computational ability.

These results have a series of implications. On one hand, they prove the relevance of accurate parameter modelling when creating predictions using these types of epidemiological models. When applying mathematical tools to find the best strategy, small changes such as adding a decay to the rate of transmission within the lockdown, as observed, affect the results drastically. On the other hand, they show the limitations of certain optimizers (ES) when applied to complex models, where they can miss the optimal solution under certain conditions. Finally, they suggest that the generalized claim that dynamical lockdowns are more efficient than continuous lockdowns is bound to the hypothesis that the rate of transmission during lockdowns instantaneously drops to negligible values. The difficulty of implementing “perfect” lockdowns can lead to situations closer to the ones we have experimented with in this work, where longer lockdowns would in effect provide better results.

## Conclusion

Over the past months, SARS-Cov2 has caused a global pandemic that has severely affected the global economy and has collapsed the healthcare systems of a large number of countries. To tackle those problems, most countries have decided to implement lockdowns without a clear strategy for when and how to implement them. In the past months, there have been two clear trends in the COVID-related studies. On the one hand, some researchers that have assessed continuous and dynamic lockdown have fixed policies in specific time frames. On the other hand, others have developed mathematical problems to decide when to implement the lockdowns. Nevertheless, we have observed a lack of studies combining both ideas. Also, we have found out that some researchers assessed the influence of a decay of the rate of transmission over time which, interestingly, resembles the curve in real scenarios.

Based on these studies, we defined the aim of this work as twofold. On the one hand, we aimed to investigate the current insight on lockdown optimization strategies, basing our analysis on a SEIR equation-based model and implementing Evolutionary Strategies (ES) as an optimizer, which differs from the commonly implemented reinforcement learning or genetic algorithm solutions. On the other hand, we aimed to explore the influence of the transmission rate parameter (*β*) on the resulting strategy proposed by the optimizer.

Those goals were assessed by considering the influence of the rate of transmission over time by setting 4 scenarios where we study combinations of constant and variable functions in and out of lockdown (On-Off, Decay-Off, On-Decay, Decay-Decay). We also set four scenarios that differed in length and number of lockdown periods, each having a total maximum number of 60 days with lockdown (12 periods of 5-day, 6 periods of 10-day, 4 periods of 15-day, 2 periods of 30-day). Furthermore, to avoid having a bias in our results towards a particular country, we assessed all the mentioned conditions on data from both Italy and Japan, which are countries that have had very different COVID transmission patterns.

Results indicated that whenever an exponential decay function is applied to the rate of transmission outside the lockdown, the optimizer shows better performance with shorter lockdowns, following a dynamic strategy and supporting the results found in the literature. However, when the decay of the rate of transmission is inside the lockdown (cases on-decay and decay-decay), the optimizer favors longer lockdowns for both countries.

Despite the fact that the applicability of these results to real cases is not immediately clear due to the simplicity of the model, this study provides a good insight of the influence of the rate of transmission’s decay when studying dynamic and continuous lockdowns, suggesting that it should be further considered in further studies with other optimizers (agent-based) and models (SEIRSHUD).

## Data Availability

Data available in a public (institutional, general or subject specific) repository that does not issue datasets with DOIs (non-mandated deposition).

https://ourworldindata.org/coronavirus

## Footnotes

1 https://graphics.reuters.com/world-coronavirus-tracker-and-maps/ COVID-19 Global Tracker, 2021

2 https://apmonitor.com/do/index.php/Main/ COVID-19Response COVID-19 Optimal Control Response, 2020

3 https://covid19-scenarios.org/about COVID-19 scenarios, 2020

## Bibliography

S. Eubank, I. Eckstrand, B. Lewis, S. Venkatramanan, M. Marathe, and C.L. Barrett. Commentary on Ferguson, et al., “Impact of Non-pharmaceutical Interventions (NPIs) to Reduce COVID-19 Mortality and Healthcare Demand”. Springer, 2020. doi: 10.1007/s11538-020-00726-x.

R. Chowdhury, K. Heng, and M.S.R. Shawon. Dynamic interventions to control COVID-19 pandemic: a multivariate prediction modelling study comparing 16 worldwide countries. Eur J Epidemiol, pages 389–399, 2020. doi: 10.1007/s10654-020-00649-w.

Julie Blackwood and Lauren Childs. An introduction to compartmental modeling for the budding infectious disease modeler. Letters in Biomathematics, 5(1):195–221, Dec. 2018. doi: 10.1080/23737867.2018.1509026.

Deanna M. Kennedy, Gustavo José Zambrano, Yiyu Wang, and Osmar Pinto Neto. Modeling the effects of intervention strategies on COVID-19 transmission dynamics. Journal of Clinical Virology, 2020. doi: 10.1016/j.jcv.2020.104440.

Thomas Rawson, Tom Brewer, Dessislava Veltcheva, Chris Huntingford, and Michael B. Bonsall. How and When to End the COVID-19 Lock-down: An Optimization Approach. Frontiers in Public Health, 8:262, 2020. ISSN 2296-2565. doi: 10.3389/fpubh.2020.00262.

Robert Glaubius, Terry Tidwell, Christopher Gill, and William D Smart. Real-time scheduling via reinforcement learning. arXiv preprint 1203.3481, 2012.

Mauricio Arango and Lyudmil Pelov. COVID-19 pandemic cyclic lock-down optimization using reinforcement learning. arXiv preprint 2009.04647, 2020.

Hado Van Hasselt, Arthur Guez, and David Silver. Deep reinforcement learning with double Q-learning. In Proceedings of the AAAI Conference on Artificial Intelligence, volume 30, 2016.

Cédric Colas, Boris Hejblum, Sébastien Rouillon, Rodolphe Thiébaut, Pierre-Yves Oudeyer, Clément Moulin-Frier, and Mélanie Prague. EpidemiOptim: A Toolbox for the Optimization of Control Policies in Epidemiological Models. arXiv preprint 2010.04452, 2020.

Harshad Khadilkar, Tanuja Ganu, and Deva P Seetharam. Optimising Lockdown Policies for Epidemic Control using Reinforcement Learning, 2020.

Luis Miralles-Pechuán, Fernando Jiménez, Hiram Ponce, and Lourdes Martínez-Villaseñor. A Deep Q-learning/genetic Algorithms Based Novel Methodology For Optimizing Covid-19 Pandemic Government Actions, 2020.

Tim Salimans, Jonathan Ho, Xi Chen, Szymon Sidor, and Ilya Sutskever. Evolution Strategies as a Scalable Alternative to Reinforcement Learning, 2017.

Darrell Whitley. A genetic algorithm tutorial. Statistics and computing, pages 65–85, 1994.

Oscar Pinto Neto, Deanna M. Kennedy, Jose Clark Reis, and Yiyu Wang. Mathematical model of COVID-19 intervention scenarios for São Paulo—Brazil. Nature Commun, 2021. doi: 10.1038/s41467-020-20687-y.

Luis Miralles-Pechuán, Fernando Jiménez, Hiram Ponce, and L. Martínez-Villaseñor. A Methodology Based on Deep Q-Learning/Genetic Algorithms for Optimizing COVID-19 Pandemic Government Actions. In Proceedings of the 29th ACM International Conference on Information Knowledge Management, CIKM ‘20, page 1135–1144, New York, NY, USA, 2020. Association for Computing Machinery. ISBN 9781450368599. doi: 10.1145/3340531.3412179.

Liu X Zhang G. Prediction and control of COVID-19 spreading based on a hybrid intelligent model. PLoS ONE, 2021. doi: 10.1371/journal.pone.0246360.

L. E. Olivier, S. Botha, and I. K. Craig. Optimized Lockdown Strategies for Curbing the Spread of COVID-19: A South African Case Study. IEEE Access, 8:205755–205765, 2020. doi: 10.1109/ACCESS.2020.3037415.

Esteban Ortiz-Ospina Max Roser, Hannah Ritchie and Joe Hasell. Coronavirus Pandemic (COVID-19). Our World in Data, 2020. https://ourworldindata.org/coronavirus.

S. R. Sukumar and J. J. Nutaro. Agent-Based vs. Equation-Based Epidemiological Models: A Model Selection Case Study. pages 74–79, 2012. doi: 10.1109/BioMedCom.2012.19.

Ottar N. Bjørnstad, Katriona Shea, Martin Krzywinski, and Naomi Altman. The SEIRS model for infectious disease dynamics. Nature Methods, 2020. doi: 10.1038/s41592-020-0856-2.

Tomas Veloz, Pedro Maldonado, Samuel Ropert, Cesar Ravello, Alejandra Barrios, Soraya Mora, Cesar Valdenegro, Tomas Villaseca, and Tomas Perez-Acle. On the interplay between mobility and hospitalization capacity during the COVID-19 pandemic: The SEIRHUD model. medRxiv, 2020. doi: 10.1101/2020.06.10.20127613.

N.M Linton, T. Kobayashi, Y. Yang, K. Hayashi, A.R. Akhmetzhanov, S. m. Jung, B. Yuan, R. Kinoshita, and H. Nishiura. Incubation Period and Other Epidemiological Characteristics of 2019 Novel Coronavirus Infections with Right Truncation: A Statistical Analysis of Publicly Available Case Data. Journal of Clinical Medicine, 2020.

S.-m. Jung, A.R. Akhmetzhanov, K. Hayashi, K. Hayashi, N.M. Linton, Y. Yang, B. Yuan, T. Kobayashi, R. Kinoshita, and H. Nishiura. Real-Time Estimation of the Risk of Death from Novel Coronavirus (COVID-19) Infection: Inference Using Exported Cases. Journal of Clinical Medicine, 2020.

L. E. Olivier and I. K. Craig. An epidemiological model for the spread of COVID-19: A South African case study, 2020. P.

Zhou, XL. Yang, and XG. Wang. A pneumonia outbreak associated with a new coronavirus of probable bat origin. Nature, 2020. doi: 10.1038/s41586-020-2012-7.

Adam J Kucharski, Timothy W Russell, Charlie Diamond, Yang Liu,, John Edmunds, Sebastian Funk, and Rosalind M Eggo. Early dynamics of transmission and control of COVID-19: a mathematical modelling study. medRxiv, 2020. doi: 10.1101/2020.01.31.20019901.

G. Giordano, F. Blanchini, R. Bruno, C. Colaneri, A. Di Filippo, Angela Di Matteo, and M. Colaneri. Modelling the COVID-19 epidemic and implementation of population-wide interventions in Italy. Nature Medicine, 2020. doi: 10.1038/s41591-020-0883-7.

Nikolaus Hansen and Andreas Ostermeier. Completely derandomized self-adaptation in evolution strategies. Evolutionary computation, 9 (2):159–195, 2001.

Frank Sehnke, Christian Osendorfer, Thomas Rückstieß, Alex Graves, Jan Peters, and Jürgen Schmidhuber. Parameter-exploring policy gradients. Neural Networks, 23(4):551–559, 2010.

Rob A Rutenbar. Simulated annealing algorithms: An overview. IEEE Circuits and Devices magazine, 5(1):19–26, 1989.

